# Exploring the gendered dimensions of health workforce retention challenges and transformative solutions in three deprived districts of Ghana: a qualitative participatory action research study

**DOI:** 10.1101/2025.03.05.25323454

**Authors:** India Hotopf, Samuel Amon, Leonard Baatiema, Patricia Akweongo, Joanna Raven

**Affiliations:** Department of International Public Health, Liverpool School of Tropical Medicine, Pembroke Place, Liverpool, L3 5QA, UK; School of Public Health, University of Ghana, P.O. Box LG 13, University of Ghana, Legon, Accra

## Abstract

The health workforce (HWF) is central to achieving Universal Health Coverage, but the ongoing global HWF retention crisis threatens progress. Women comprise 70% of the HWF and yet face unique retention challenges. Gender transformative actions on HWF are needed, but there is a knowledge gap, especially in low-resource settings. Ghana is facing a HWF crisis in deprived, remote areas. A project that co-designed and piloted retention interventions in three such districts highlighted gendered dimensions. This study sought to elucidate the gendered dimensions of HWF challenges and make gender transformative recommendations.

This qualitative study embedded a participatory action research and intersectional approach. Thirty-six (36) key informant interviews were conducted to explore retention challenges, current policies/activities and recommendations. Respondents were purposively selected for cadre and gender, striving for equal geographical distribution. Intersectional gender analysis was conducted using the framework analysis approach and Morgan’s gender framework.

Women health workers dominated deprived districts, with the small number of men assigned to island communities, as men were deemed more resilient to difficult conditions. There was a shortage of women health workers, and retention was low, primarily due to family responsibilities. Most women juggled difficult working conditions with singlehanded childcare and responsibility for maintaining relationships, hindered by poor telecommunication networks and transport challenges. Many women feared boat crossings and cannot ride motorbikes, necessitating expensive motorbike rider hire, with some attributing long journeys to miscarriages. Women face expensive accommodation, barely covered by salaries – this, combined with high food costs and inability to conduct locum work, causes financial stress. Safety and security concerns, including robberies, motorbike accidents and sexual harassment were commonly highlighted. Current policies are not gender transformative and failed to address women’s challenges. Recommendations include tailoring incentives to women with childcaring responsibilities, investing in accommodation and security, strengthening community support, sexual harassment policy and awareness.

## 1. Introduction

The HWF plays an integral role in in the overall function of the health system and in ensuring equitable health care service access (1). HWF retention remains a global challenge, particularly in low-and middle-income countries, including the African Region, where the disease burden is high and health systems are typically under-resourced, with the density of health workers (HWs) being particularly low in remote areas (1–4). Addressing HWF retention is key in realising Sustainable Development Goal (SDG) 3 Good health and wellbeing, which includes achieving Universal Health Coverage (UHC) (1, 5).

Sub-Saharan Africa has approximately 2.5 HWs per 1,000 population; well below the World Health Organization (WHOs) recommended SDG density threshold of 4.45 per 1,000 population (6, 7). Ghana is experiencing a HWF crisis, characterised by the mass exodus of HWs and difficulty attracting and retaining HWs in deprived, remote areas, particularly among nurses (8). Whilst the Ghanaian Health Service (GHS) estimated a national HWF density of 2.65 per 1,000 population in 2017, the WHO estimated a density of 2.22 per 1,000 population (9–11). There is spatial variation in HWF density nationally, with a lower density in hard-to-reach, deprived districts, characterised by poor road networks, poverty and a lower density of health facilities (HFs) (8). Whilst Ghana has made considerable strides towards gender equality compared with other countries in the West and Central African Region, women and girls continue to experience an unequal stance in society, marked by women accounting for just 15% of the national parliament (12, 13). Aside from negatively impacting health outcomes and quality of care, the ongoing retention crisis risks SDG progress (14).

Gender is defined as “the socially constructed roles, behaviours, activities, attributes and opportunities that any society considers appropriate for men and women, boys and girls” and people with non-binary identities (15, 16). Globally, women make up ∼70% of the HWF (17). Gender norms and power relations shape the development, attraction, recruitment and retention of HWs, with women disproportionately impacted by certain challenges e.g., sexual harassment, discrimination, family responsibilities and a global 24% gender pay gap, affecting retention (17–19). Hence, understanding and addressing gender inequities is central in addressing the retention crisis. However, a gap in gendered HWF data and literature remains, particularly in LMICs, where the challenges are greatest (17). Evidence is needed to enable the implementation of gender transformative interventions to strengthen the retention of women HWs (17).

To address the ongoing HWF retention crisis, the University of Ghana School of Public Health and the Liverpool School of Tropical Medicine launched a participatory action research project to co-design, implement and evaluate community-based interventions to strengthen HWF retention in three most deprived Ghanaian districts. Findings from the situational analysis, a review of Ghana’s policies related to retention (Table 1) and participatory workshops highlighted stark gender inequities, influencing retention.

**Table 1.**
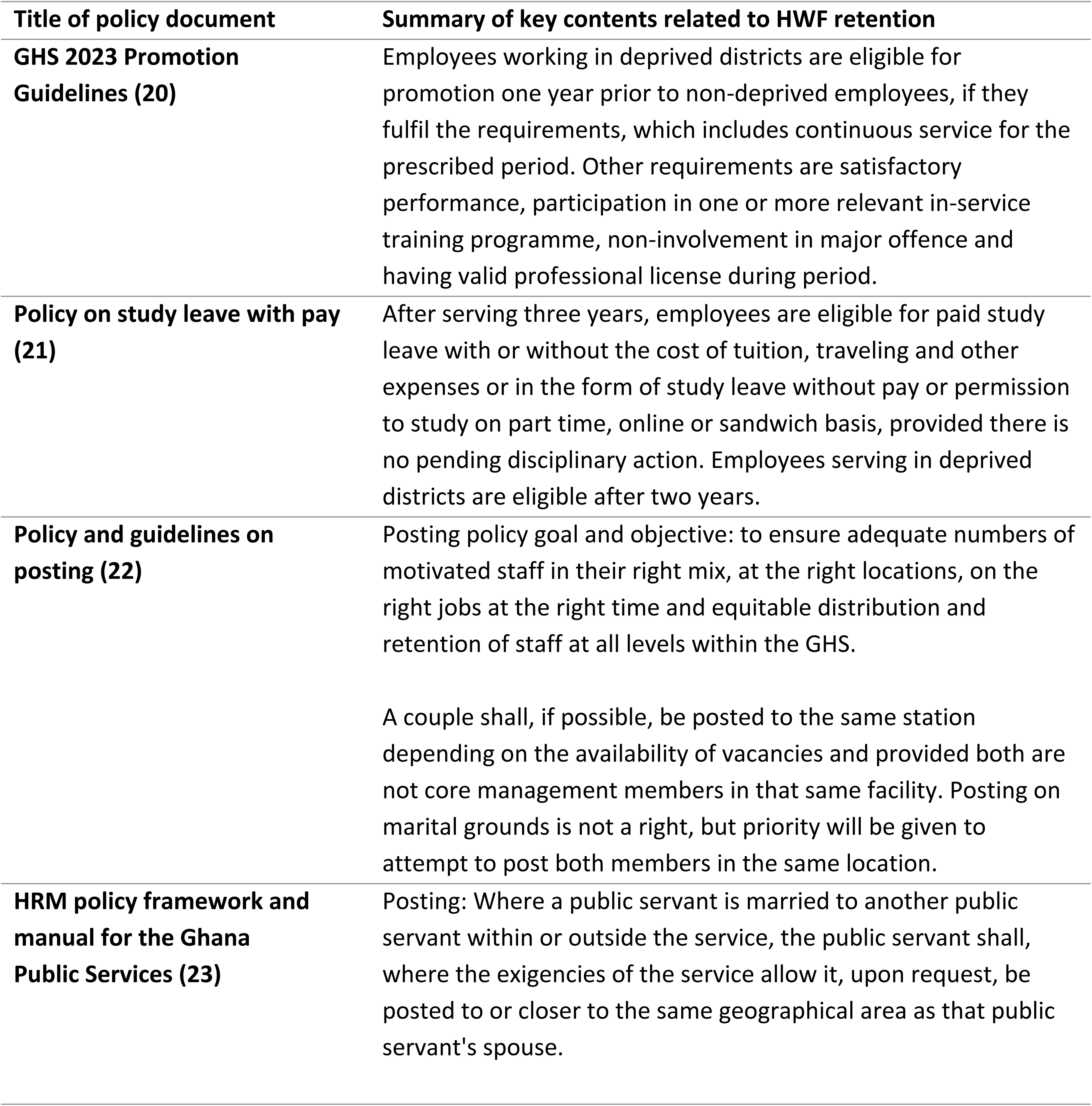
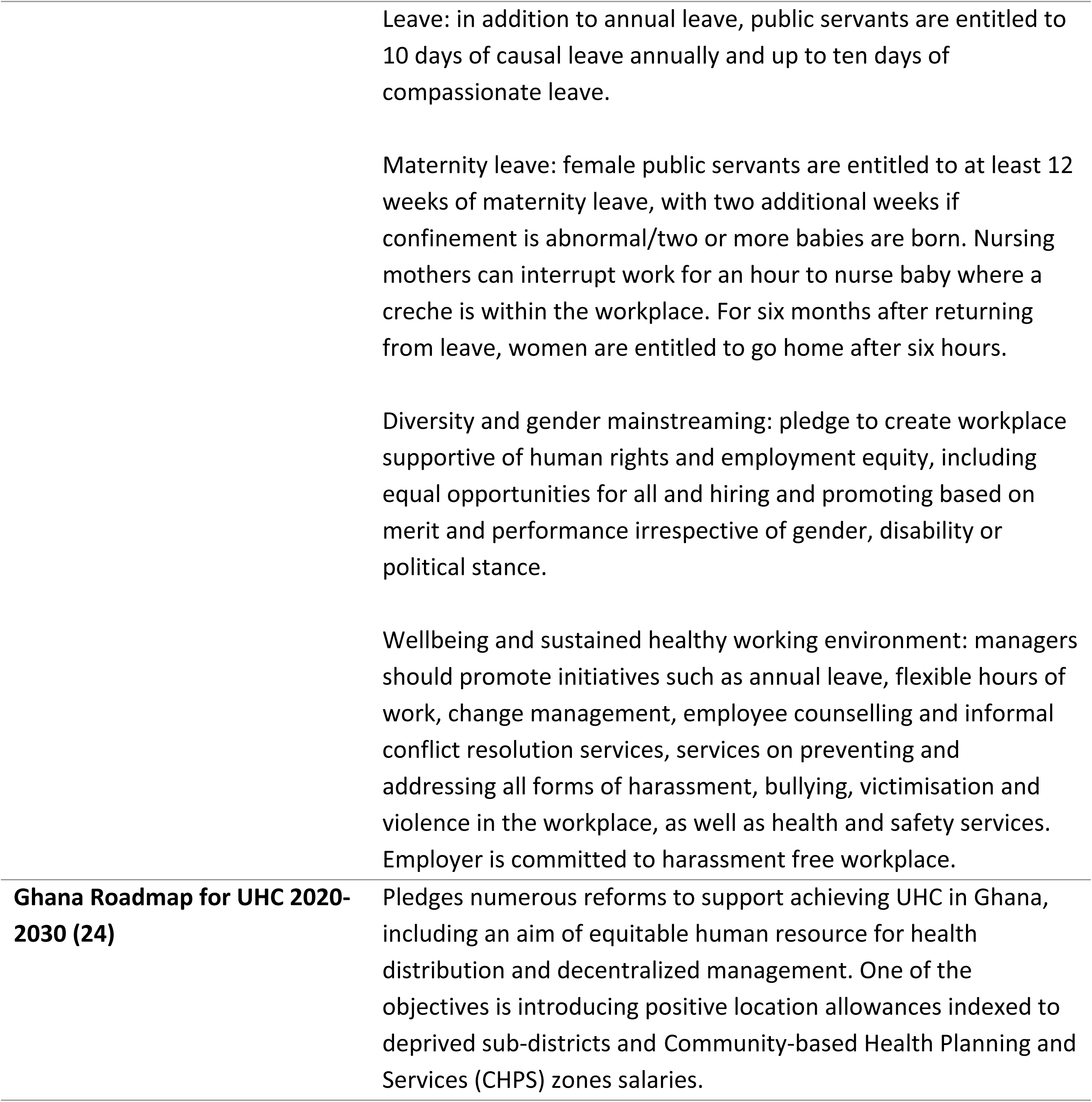
Summary of key policies related to HWF retention and gender.

| **Title of policy document** | **Summary of key contents related to HWF retention** |
| --- | --- |
| **GHS 2023 Promotion Guidelines (20)** | Employees working in deprived districts are eligible for promotion one year prior to non-deprived employees, if they fulfil the requirements, which includes continuous service for the prescribed period. Other requirements are satisfactory performance, participation in one or more relevant in-service training programme, non-involvement in major offence and having valid professional license during period. |
| **Policy on study leave with pay (21)** | After serving three years, employees are eligible for paid study leave with or without the cost of tuition, traveling and other expenses or in the form of study leave without pay or permission to study on part time, online or sandwich basis, provided there is no pending disciplinary action. Employees serving in deprived districts are eligible after two years. |
| **Policy and guidelines on posting (22)** | Posting policy goal and objective: to ensure adequate numbers of motivated staff in their right mix, at the right locations, on the right jobs at the right time and equitable distribution and retention of staff at all levels within the GHS.<br>A couple shall, if possible, be posted to the same station depending on the availability of vacancies and provided both are not core management members in that same facility. Posting on marital grounds is not a right, but priority will be given to attempt to post both members in the same location. |
| **HRM policy framework and manual for the Ghana Public Services (23)** | Posting: Where a public servant is married to another public servant within or outside the service, the public servant shall, where the exigencies of the service allow it, upon request, be posted to or closer to the same geographical area as that public servant's spouse. |

Women account for more than 70% of Ghana’s HWF, but are overrepresented at the lower end of the health system, wherein just 35% are medical doctors and women constitute almost 86% of nurses and midwives (20). Whilst there is some literature on HWF retention challenges in Ghana (21–23), to our knowledge, there are no qualitative studies exploring the gendered dimensions of retention challenges within deprived districts, where retention challenges are greatest.

Therefore, this study sought to elucidate the gendered dimension of HWF challenges in the three most deprived Ghanaian districts and present evidence-based recommendations, through 1) describing differences in HWF retention challenges between women and men and 2) making recommendations for gender transformative actions. The study will contribute to the limited evidence-base, ultimately supporting the design and implementation of gender transformative HWF strengthening interventions.

## 2. Materials and methods

### 2.1 Study design

This qualitative study draws on findings from the situational analysis stage of a mixed-methods participatory action research (PAR) study which aims to co-design, pilot and evaluate HWF retention strengthening interventions in the same setting. A PAR approach was used to ensure local ownership and participation in decision-making, meaningful equitable participation from different social groups and intervention sustainability.

To explore gender in relation with other overlapping social stratifies, we applied an intersectional lens throughout – specifically, the intersectionality wheel and gender analysis domains derived from Morgan, George (24) (25, 26). This is critical to ensure health systems consider and address the multiple intersecting inequalities faced by HWs and patients instead of exploring individual forms of oppression (16, 27).

### 2.2 Study setting

The study was conducted in the most deprived districts: Kwahu Afram Plains North (KAPN), Kwahu Afram Plains South (KAPS) and South, and Kwahu East (KE). Our situational analysis report (unpublished) demonstrated that districts are characterised by high unfilled posts (36-50% posted but refuse postings), low cadres and high doctor-to-patient ratios (e.g., one doctor: 70,000 populations in KAPN), resulting in poor health outcomes. The districts have a population of 70,837 (KAPN), 78,974 (KAPS) and 83110 (KE), respectively. Whilst all three districts have the same number of health centres (5), there is variation in the number of CHPS Compounds, ranging from 23 in KE to 39 in KAPN, which also has the only hospital. All districts have high rates of multidimensional poverty, with the lowest rate of 29.4% in KE (KAPS = 39.1%; KAPN = 48.5% versus national average of 24.3%) – potentially due to KE being inland, whilst KAPS and KAPN are remote islands (28).

Districts are managed by a five-core member team, called District Health Management Team (DHMT), with the districts divided into 28 sub-districts, managed by sub-district heads, managing health centres. Each sub-district has several CHPS compounds providing basic primary and community health services. There are 10 sub-districts in KAPN and nine each in KAPS and KE, all of which were included in the study.

### 2.3 Participants and sampling

Key informants were purposively sampled from across three health system levels: district health directorate (DHD), sub-district and district hospital (DH), with a mix of frontline HWs and managers sampled. Additionally, community members involved in local level health services management were recruited. Participants were eligible if current health workers or community members of target districts, aged 18 years or older and provided informed consent.

Participants were purposively selected with maximum variation for participant type and gender, striving for an equal distribution across the three districts (Table 2). A sampling frame was obtained from the DHMT and a convenient sampling approach was used. Informants were randomly selected, stratifying for district and job role to ensure maximum variation. Recruitment was coordinated via the national, regional, district, and sub-district heads informants and consenting participants were contacted via phone call.

**Table 2.** KII sampling matrix. Note that there are no district hospitals in KAPN or KE, hence there are 0 district hospital informants.

| District | Health system level | Women, N (%) | Men, N (%) | Total, N (%) |
| --- | --- | --- | --- | --- |
| KAPS | DHD | 0 (0%) | 3 (100%) | 3 (100%) |
|  | Sub-district | 5 (100%) | 0 (0%) | 5 (100%) |
|  | Community | 0 (0%) | 2 (100%) | 2 (100%) |
|  | DH | 1 (50%) | 1 (50%) | 2 (100%) |
| KAPN | DHD | 1 (25%) | 3 (75%) | 4 (100%) |
|  | Sub-district | 5 (83.3%) | 1 (16.7%) | 6 (100%) |
|  | Community | 0 (0%) | 2 (100%) | 2 (100%) |
|  | DH | 0 (0%) | 0 (0%) | 0 (0%) |
| KE | DHD | 0 (0%) | 5 (100%) | 5 (100%) |
|  | Sub-district | 5 (83.3%) | 1 (16.7%) | 6 (100%) |
|  | Community | 0 (0%) | 1 (100%) | 1 (100%) |
|  | DH | 0 (0%) | 0 (0%) | 0 (0%) |
| **Total** |  | **17** | **19** | **36** |

### 2.4 Data collection

The first 21 KIIs were conducted between February – April 2024, in person during district visits. As there was a low number of female key informants included, a second round was conducted in July – October 2024 with more women participating. Six were conducted in-person with sub-district managers at a reflection workshop and the remaining nine were conducted via telephone calls. Topic guides were informed by the HWF challenge literature and explored: the current HWF retention situation; determinants and impacts of retention challenges; existing approaches to address challenges and suggestions. This included questions on the difference in challenges among men and women and whether policies and guidelines were gender tailored. Additional interviews with women HWs included questions on sexual harassment, highlighted as an issue in participatory workshops, as well as avenues for gender transformative actions. KIIs were conducted by core members of the research team who were familiar with the study tools and had knowledge and experience in conducting qualitative research. All interviews were conducted in English and audio recorded, with field notes also taken.

### 2.5 Analysis

Audio recordings were transcribed verbatim and quality checked, and all identifiable information was removed, with transcripts assigned unique codes and uploaded to SharePoint – only accessible to core researchers. IH conducted intersectional gender analysis using the framework approach, with support from NVivo 12, which included gender disaggregating data (29). A deductive framework based on the topic guides was used initially, with inductive themes iteratively incorporated. Next, matrices were exported in Excel and data was charted in Word, with challenges categorised using the gender framework from Morgan, George (24). Throughout, similarities and differences were drawn across districts, health system levels and gender.

### 2.5 Ethical considerations

Ethical clearance was obtained from the Ghana Health Service (GHS-ERC:007/02/24) and LSTM (24–002). All participants were adults who written informed consent, with the voluntary nature of participation emphasised. Confidentiality was maintained through removing identifiable information, conducting KIIs in private locations and storing anonymized data in a secure location, with restricted access. Refreshments and reimbursement for travel costs were provided to all participants where necessary.

## 3. Results

### 3.1 Perceptions of gender and health workforce situation

Across districts, women and men consistently reported that women account for most of the workforce, with respondents in KAPN emphasising that men are sent to islands, with just one woman posted. Whilst most women and men across districts deemed female retention low, in KAPS, only sub-district managers and HWs perceived gendered differences.

### 3.2 Gendered drivers of attrition

Below we explore the unique challenges faced by women and men living and working in deprived districts, driving attrition. Challenges are explored through the gender analysis dimensions described by Morgan, George (24).

**Figure 1.**
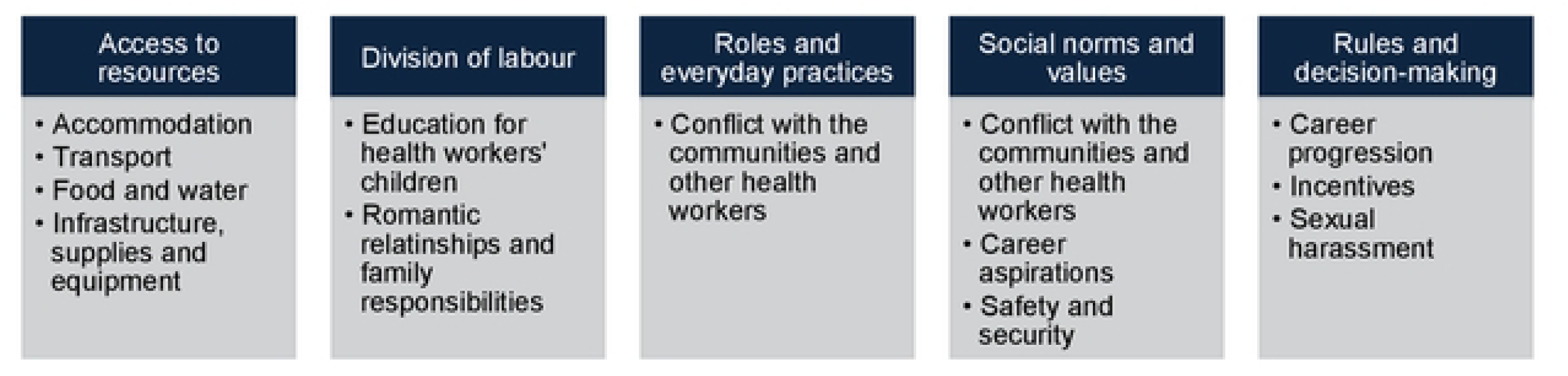
Overview of health workforce challenges explored according to Morgan et al’s (2016) framework.

#### Access to resources

##### Accommodation

All women described accommodation issues, with many deeming women disproportionately impacted. In KAPN, women commonly highlighted a lack of accommodation, with one reportedly combining the storage and pharmacy HF rooms to create a bedroom. Where available, cost of rent is high compared to Accra, driving posting rejections/withdrawals and perceptions of punishment. Additionally, KAPS women highlighted poor infrastructure (e.g., “mud houses”), with one mentioning many share rooms, impeding them from accommodating mothers for childcare support, and one described having to refuse to share with a male colleague as she felt uncomfortable. Some described animal infestations, and long commutes, whilst one noted that men on islands lack toilets, necessitating bathing outside. All KE women highlighted similar issues, with one HW recalling new staff pleading for transfers, and a manager stating that HWs must relieve themselves in the bush due to mud huts lacking toilets, – consequently, retention is low, and fewer staff are posted (e.g., three versus five).

Generally, men perceived accommodation as less pressing, especially in KAPS. In KAPN, most reported staff accommodation shortages, necessitating renting further afield, introducing financial and logistical issues, with accommodation commonly lacking basic amenities. In KAPS, some highlighted absence of televisions, high rent and infestations. Similarly, most KE men highlighted a lack of televisions, water and poor infrastructure, with some DDs respondents attributing the absence of free accommodation to attrition.

##### Transport

Transport was a major challenge for men and women, with women’s unique challenges with motorbikes and boat crossings reportedly hindering resilience. Conversely, men are perceived as “tough” and resilient to island conditions, with a few KAPS women asserting such stereotypes disadvantage men. Most KAPN women highlighted challenges, including fearing motorbikes/boat use, especially during rainy season or pregnancy, necessitating expensive “okadas” (motorbike taxis). This drives refusals/withdrawals, perceptions of being punished, and male island postings. All KAPS women echoed these sentiments, with some reporting that okadas constitute most of their salaries and fear boats if menstruating or with children, though one recalled women HWs jointly purchasing and learning to ride motorbikes. In KE, most women feared motorbike usage, especially during rainy season, with long motorbike journeys attributed to miscarriages by several women, with one calling for more male HWs as pregnant women cannot ride.

> “Y*ou people can get an accident. When it happens like that, your leg can break, and you cater for yourself. Nobody will even assist you. So, all those things are hindrances. And at times, you can go and meet some people, they will attack you, trying to come and rape you on the way.” (Female SD HW, KAPN)*

All KAPN men described challenges, including boats/cars/motorbikes shortages and psychological/safety issues due to boats and poor roads, with some highlighting women’s boat/motorbike fears and unique safety issues. Most KAPS men echoed sentiments, emphasising financial/time costs and women’s difficulties, owed to KAPS’s poor “advertising image”, (e.g., “scary place to work” or “trapped”) by some, driving rejections and attrition. KE echoed challenges, with some emphasising women’s motorbike discomfort, punishment perceptions and okadas charging twice at night, hindering evening class access, mentioned by an SD.

##### Food and water

Food and water issues were common among women compared to men, especially in KAPN, where most women reported paying for imported water and some highlighted insufficient community support, with one exception. Additionally, a few KAPN women described unpalatable, expensive food. In KAPS, a few women reported purchasing imported water and whilst some praised the food, including a woman whose community helped her start farming, some faced difficulties obtaining food, especially on islands. Only a few KE women expressed difficulties accessing potable water.

> *“They will sell it to you for two cedis because they feel that you are a government worker, and you have money. Yeah. So, things that they are supposed to sell at their normal subsidized price, they make it something else […] But initially, that wasn’t how things were […] You’d be there, and they would bring you foodstuff. They would come to clean your place for you, come to fetch water for you. Like they want you to feel that kind of belongingness.” (Female SD manager, KAPN)*

Fewer men faced challenges; in KAPN, some highlighted being overcharged for food/water and another noted assemblyman’s wives cook for male HWs only on islands, because “in some peoples’ cultures, men don’t cook”. Issues with food/water unavailability/pricing (including fish unless fishing based in fishing communities) on islands was highlighted by a few KAPS men, while some KE men, including DDs respondents and HWs, reported potable water shortages.

##### Network and electricity

Many women highlighted challenges, compared with men. In KAPN, 50% highlighted network and electricity issues, deemed the greatest female attrition driver by a sub-district manager, though one HW felt men on islands are most impacted and another reported good network/electricity access. This was echoed by half the KAPS women, with some emphasising disproportionate impacts on women, due to family disconnect and some describing negative service delivery impacts. Similar relationship and service delivery challenges were described by most KE respondents, with a common sentiment of having to “choose either your work or relationship”.

> *“If the partner comes, there’s no network there. Why will I come to a place where there’s no work? My work people call me, so the partner also. I can’t continue with this, so I have to break up because I cannot engage with somebody who is nowhere. […] it depends on you, the individual to choose either your work or your relationship. […] It is one of the things that is very hard as women, but we love and have passion for the job.” (Female SD manager, KE)*

A few KAPN men described island electricity/network challenges, hindering communication with families. A couple of KAPS CMs and many HWs described electricity/network issues, impacting service delivery (e.g., contacting field staff), with a KE manager also emphasising disproportionate impacts on women. However, a couple of men in KAPN and KAPS described grids/solar panel installation addressing challenges.

##### Infrastructure, supplies and equipment

In KAPN, most women described issues with HF infrastructure (e.g., bats, leaks, insufficient beds and privacy), echoed by some KAPS women, with one reportedly having to work under a tree. KE women commonly highlighted supplies and equipment shortages, with some reporting insufficient inpatient beds. Across the districts, most men echoed similar infrastructure issues, with one man in KAPN attributing inadequate infrastructure to female attrition, whilst some men in the other districts also emphasised equipment shortages.

#### Division of labour, roles and everyday practices

##### Education for health workers’ children

Except for KAPS, few women mentioned education, conversely, most men highlighted education for HWs children as a key female attrition driver. In KAPN, a few women felt education drove female attrition, though one noted the recent introduction of a creche, with just one sub-district manager in KE mentioning education challenges. Education was more prominent in KAPS, owed to women’s attrition/posting refusals by most, with several women having sent children to live in Accra for better education.

In KAPN, a few men reported that women leave and join their husbands due to children’s education, whilst most men in KAPS and KE echoed the sentiment. A few men in KE described a lack of day care, making it difficult to for them to reject women’s transfer requests, with one DH worker stating that women must bring babies to meetings. However, a couple of KAPN and KAPS men reportedly grappled with decisions regarding their children’s education.

##### Romantic relationships and family responsibilities

Romantic relationships and family responsibilities were deemed key challenges by all women and most men, with women HWs bearing the brunt of responsibility. Across districts, women commonly grappled with juggling careers and relationships, both in terms of maintaining relationships and finding partners to start families with, typically preferring to find partners outside of communities where they work, due to a lack of suitors. With a few exceptions, women bared responsibility of travelling back and forth during annual leave/weekends maintaining communication with partners, with fear of boats/motorbikes and menstruation impeding efforts. Conversely, men typically brought girlfriends/wives with them or found partners in communities. Some women felt the associated stress, combined with long motorbike journeys, caused miscarriages, with one KAPN woman experiencing multiple miscarriages without her husband’s support. She also emphasised issues with loyalty and trust, adding that in Ghana men typically pay dowry.

> *“No, he doesn’t live with me. So, I have to travel all the way to the Volta region to meet up with him. That’s one problem. According to our Ghanaian culture, you are to live with your husband who has paid your dowry. But because of working conditions, you’re not getting that. Sometimes it’s challenging. When I had my issues (multiple miscarriages), I wanted comfort but because he was often not around to be a source of comfort for me at that moment; later on, he came through. But at that moment, I had to embrace everything by myself. So, it’s equally a challenge. If your husband is not the understanding type, your marriage life will not be stable. It can bring a lot. There may be mistrust, there may be an issue with loyalty, and blah blah. It will all make the marriage not work. […] And as a woman, there’s a time limit for us.” (Female SD HW, KAPN)*

Consequently, most women only see their partners a few times a year, raising their children without support from husbands or family members, with insufficient feeding rooms and creches exacerbating challenges. Some women also described having to travel home regularly to care for sick parents. Across the board, women consistently highlighted these challenges as one of the greatest attrition drivers.

> *“As a woman too, there are some places that, maybe if you are breastfeeding your child, you don’t get a place to do that, do you understand? But if you are in town or a bigger facility, they will create a place that maybe you can feed your child. You feed your child in a place like Atibie; it is an example I am setting. They have a place for their school children. So, when you give birth, you send your child to their school, even toddlers, to take care of your child.” (Female SD manager, KE)*

All KAPN men, and most in KAPS and KE echoed this sentiment, with some emphasising the added pressure on women attempting to start families. Many highlighted that “women don’t usually have much say” in initiating relationships with community men and some asserted that only “outstanding” husbands/partners would visit partners in deprived districts. A few men also emphasised women’s caring responsibilities for sick parents, though one DHD respondent in KAPS emphasised men’s responsibility and one was reportedly caring for his parents. Ultimately, most men attributed attrition to women wanting to join their partners.

#### Social norms and values

##### Conflict with the communities and other HWs

Women experienced community conflict more commonly, and whilst men consistently highlighted conflict between women HWs, women rarely highlighted the challenge. Some KAPN women reported insufficient community support in improving facilities, though one asserted the community were “doing their best”. Communities were deemed demanding, unsupportive, and in one case, financially extortionate by some KAPS women, with a few exceptions. Others recalled community leaders commandeering HF rooms and communities complaining upon HWs attending conferences. Community conflict was common in KE, with some requesting durbars to discuss issues with HWs and asserting a lack of respect for nurses. Whilst one man perceived his community as supportive, a few described issues such as community members stealing chickens from HWs, and insufficient HW support as HWs are salaried “government workers”. One man concluded that unless communities are “safe place” you will be unhappy.

In KAPN, only the sub-district manager described conflict between staff, involving male colleagues, while a few sub-district managers in KAPS and KE also highlighted conflict between staff. This included some describing “petty quarrels”, primarily among women. However, none of the women HWs described issues. Among men, a DHD in KAPN asserted the inevitability of conflict, especially among women, while just one KAPS man highlighted conflict generally. In KE, most men attributed conflict between women to attrition, due to “petty issues” (e.g., unclean bathrooms), though a DHD stated that some men also “misbehave” with management if unhappy with their posting.

##### Safety and security

Safety and security was a key challenge for all, primarily in KAPN and KAPS. Most KAPN women described issues, including unsafe bridges, infestations and numerous incidents, including a patient crashing his motorcycle into a woman’s accommodation when she refused to open her door at night. Others described communities setting HWs cars alight and robberies/rape/accidents during motorcycle outreach, despite reported declines in crime. In KAPS, many women echoed sentiments, typically feeling unable to protect themselves during independent outreach, compared with men. A sub-district manager highlighted pregnant women must also ride motorbikes, recalling a woman nearly dying in an accident and only receiving one weeks leave. Some women emphasised living alone, with no security, and one reported animal infestations. In KE, some women also highlighted motorbike accidents and insufficient security and lighting which drives fear of late-night patient visits, though one felt security had been addressed.

> *“If something is happening to you and let’s say you’re screaming, no one will listen here. There is no security and just imagine I’m alone. So sometimes when someone comes to knock on my door, let’s say at dawn, I don’t go out because I’m scared […] But maybe the person is not feeling well, yes. But if you come and knock at my door, you wouldn’t expect that. Because where I am, after the building there, the places you see surrounding me are forests and cemeteries. So, you wouldn’t expect me to come out. Even if I recognize the voice, I can’t come out because I’m scared. Yet I may need to be able to attend to the person at that particular moment.” (Female SD HW, KAPS)*

Most KAPS men highlighted transport safety issues (e.g., lack of life jackets, insurance and delayed boat referrals resulting in HWs death), whilst a DHD was reportedly shot during an attempted robbery, and another asserted security is driving female attrition. Despite one man reporting a crime decline, most KAPS men had experienced or heard of shootings and robberies, with one nearly dying. Some attributed safety/security challenges (e.g., nighttime patient visits, motorbike accidents, life jackets and insurance) to female attrition, also echoed by some KE men.

##### Career aspirations

Some respondents, primarily women, felt women were better suited to nursing and midwifery due to more empathy and compassion, making patients feel more comfortable, with one noting that her midwifery school stopped enrolling men recently, highlighting gender stereotypes. Some women attributed the lack of male HWs to men being better suited to shorter working hours, acute hospital roles or non-medical roles like teachers or accountants, due to men’s fear of blood. A woman sub-district manager lamented that even male nurses are referred to as doctors, illustrating the gender inequities. Whilst one man asserted that “You don’t need to be a woman to be a nurse”, some men in KAPN asserted that patients become “shy” and may reject care from male midwives.

#### Rules and decision-making

##### Career progression and income generation

Whilst many women in KE had negative experiences, the study leave, and promotion policies were praised in KAPN and KAPS. However, men consistently highlighted career progression challenges, especially for women HWs in KE. In KAPN, just one sub-district manager reportedly benefitted from study leave – the main factor in her willingness to stay in the district. Linked to education, one respondent recalled being mocked by urban colleagues for speaking local dialects. Most women in KAPS praised study leave, though one woman’s colleague was rejected repeatedly over a decade. Most KE women deemed the study leave policy ineffective, with numerous accounts of serving 6-8 years without leave, due to unsupportive directors and staff shortages. Some emphasised being unable to attend weekend/evening classes.

> *“Your English will not be all that good because you have not been speaking English. […] So, whenever we go for workshops, we feel bad. Are we also nurses at all? Because we are from the village, so for some people, it normally creates an impact on them. […] The moment you appear, those in cities, they try to mock and say that “oh, the villagers have come. They have come. The villagers, they have come.” So, it’s not encouraging us to stay back because there is a director who is trying to mock.” (Female SD HW, KAPN)*

In KAPS, some men described issues with promotion policies, and most deemed the study leave unsuccessful, whilst some emphasised that weekend study is “extremely expensive” and logistically challenging. This contributes to perceptions of deprived HWs “wasting their time”. One CM asserted that aside from joining their husbands, most women leave for education. All men in KE echoed study leave sentiments, with one emphasising that staff must use limited annual leave to sit exams, and another attributing study leave challenges to attrition in women.

A couple of KAPS women, including a sub-district manager, asserted that promotions-out-of-turn and exposure to greater activities/responsibilities supports progression, though another HW deemed the promotion process expensive, time consuming and unfair, reportedly failing having refused to bribe staff. In KE, most women expressed frustration with career progression, with some highlighting men leaving for greener pastures with kinder working hours and conditions. However, a psychiatric nurse felt enlightened due to greater exposure to activities (e.g., supporting deliveries) and responsibility, reportedly benefitting her career.

Most men in KAPN attributed male attrition to seeking “greener pastures”, with some deeming the promotion policy ineffective. Some KAPS men described issues with promotion policies.

Regarding income generation, many women, primarily in KAPN, emphasised their inability to conduct locum work as a major challenge compared with urban colleagues, especially considering the high cost of living.

##### Insufficient incentives

Across districts and cadres, women consistently highlighted insufficient financial incentives, whilst aside from one individual in KAPS, only men in KAPN described challenges. In KAPN, half of the women emphasised the high cost of living compared to financial incentives, with a sub-district manager and DHD worker emphasising that HWs “struggle to make ends meet” and another emphasising that whilst there are deprived financial incentives, staff are owed backpay. Most KAPS women highlighted insufficient financial incentives, inability to conduct locum work and high out of pocket costs (e.g. hiring translators), with one emphasising that despite discussions, rural incentives remain absent. Another HW feared reshuffling, wherein you are moved “without reason”, incurring stress and financial costs. Most women in KE echoed sentiments, emphasising the high cost of living, with two reportedly having no money at the months end. Moreover, a couple of women described paying out of pocket for workshops and referrals. In KAPN and KAPS, some men, primarily DDs and DHs, emphasised inadequate financial incentives considering the high cost of living, with some “struggling to make ends meet”, driving attrition. A DHD respondent in KAPN also implied issues with financial misconduct and a DH worker in KAPS recalled regional colleagues joking that they would refuse deprived postings, even if offered 10,000 cedis/month, highlighting the negative perceptions.

Some respondents described insufficient non-financial incentives, primarily in KAPN. In KAPN, some women emphasised a lack of health insurance despite risks (namely women’s transport), whilst a sub-district manager highlighted insufficient financial support and persistent negative feedback on revenue generation. A couple of KAPS women highlighted inadequate healthcare access, in terms of hospital treatment charges, and HWs “walking sick” due to harsh working conditions and insufficient care, with one HW discovering he had cancer and dying, due to inadequate access. One sub-district manager reportedly provided encouragement and paid out of pocket for staff incentives (e.g., food/drink/fuel), though other managers were unaware of incentives and no KE women discussed them. Some KAPN men highlighted the lack of insurance despite safety challenges, and one emphasised his district only recently being categorised as deprived, whilst a DHD man asserted that managers typically overpromise during recruitment, driving attrition. A couple of KE men emphasised KE not being categorised as deprived, denying HWs beneficial policies, reportedly driving posting refusals.

##### Sexual harassment

During the second interview round, women were asked about sexual harassment and related policies. Overall, sexual harassment appears prevalent within communities in KAPS and KAPN, with just one workplace incident.

Most KAPN women described incidents in the community and bush, including rape attempts and sexual propositions for amenities, with one attributing violence to farming communities where “young ones don’t have much to do” due to unemployment. Some women were unaware of incidents, with one asserting the prevalence is higher on islands and incidents are hidden, due to stigma and insufficient confidentiality. Community incidents, including attempted rape, harassment and repeated proposals, were described by most KAPS women, with one asserting communities perceive HWs as “someone special”. Just one sub-district manager described a workplace incident, wherein a woman HW was pressured into accepting a colleague’s proposal, due to safety fears, exacerbated by her remote, insecure setting, with the man eventually becoming physically violent. She lamented over the woman not speaking up, emphasising sexual harassment as a hidden issue which is worse on islands. Conversely, most KE women were unaware of incidents, though some described local men using “juju” (magical powers) to seduce HWs, emphasising women’s isolation, and one recalled community members visiting facilities at night, saying “things” and trying to “hold you”. Another HW was apprehensive to speak on the subject, since explaining that it is difficult to know if accounts are truthful and evidence may be requested.

> *“There were two CHNs, but one was a senior colleague, a man. And then the lady came. So, when the lady came, it was like he proposed to the lady. The lady didn’t want to accept. […] But the lady also got scared because the moment you want to report an issue; it would be like she’s reporting the in-charge. So, she kept it to herself. […] The lady really got scared because where they were, it was not closer to the town. They are in the town and where they are, there is no light. It’s the solar that is only meant for the facility. This lady had no one to talk to, so she had to comply […] the guy thought she had another boyfriend. And then told her to bring the phone and the girl refused. So, he laid his hands on her and people around came to the rescue of the lady.” (Female SD manager, KAPS)*

Whilst some KAPN women described informal responses (e.g., advising women not to conduct outreach alone and warnings of penalization from community chiefs), only one HW was aware of formal policies – collaborating with social welfare for report and follow-up of cases, which she deemed effective. None were aware of formal policies in KAPS, though informal actions included community members beating a perpetrator and one sub-district manager ceasing to post pairs of women and men together. Similarly, no KE women were aware of formal policies and whilst one described community punishment (community service), she emphasised the health system and chief’s failure to address “juju” marriages.

### 3.3 Current practice and recommendations for gender tailored approaches to addressing retention

Across districts, no women were aware of gender tailored support, with many asserting men and women should be treated equally – however, KAPN and KAPS participants reported some informal support. In KAPN, one sub-district manager believed in equal treatment, but noted women are rarely posted on islands, whilst the other manager reportedly allowed older unmarried women additional leave to visit home and find partners. In KAPS a HW noted that women are rarely sent to islands.

None of the men reported formal tailoring, though there were several informal examples. In KAPN, a few highlighted that women are favoured when granting annual leave, especially if pregnant, whilst a couple of DD respondents in KAPS reported an emphasis on one-to-one support for women, especially single mothers. One man reported providing additional support completing transfer and career progression forms, due to women’s poorer IT skills and one individual asserted that men are disadvantaged, due to additional informal support provided to women. In KE, a sub-district manager reportedly avoids placing mothers/pregnant women on night shifts, as “children need to be with their mothers”, adding that there is an emphasis on appreciating women HWs, especially midwives, during award ceremonies.

When asked about gender transformative support for women HWs (Table 3), women commonly called for improved access to affordable, secure accommodation, with one calling for gender segregation. This was followed by requests for better access to education and reducing the necessity for women to ride motorbikes, especially when pregnant, as well as improving financial support for women HWs in deprived districts. Men were not explicitly asked about gender transformative support, though five KAPS and KE men made suggestions, related to security and conflict resolution.

**Table 3.** Summary of HWs suggestions for gender transformative actions to strengthen retention.

| Recommendation | Women |  |  |  | Men |  |  | Both |  |
| --- | --- | --- | --- | --- | --- | --- | --- | --- | --- |
|  | KAPN | KAPS | KE | Total | KAPN | KAPS | KE | Total | Total |
| Work with communities to provide affordable, secure, gender segregated accommodation | 3 | 4 | 4 | 11 | - | - | 1 | 1 | 12 |
| Improve access to staff education and training | 5 | 2 | 0 | 7 | - | - | - | 0 | 7 |
| Strengthen managerial communication and support | 1 | 2 | 1 | 4 | - | - | 1 | 1 | 5 |
| Improve HF infrastructure and equipment | 1 | 1 | 1 | 3 | - | - | 2 | 2 | 5 |
| Reduce women's motorbike outreach when pregnant and or alone | 3 | 0 | 1 | 4 | - | - | - | 0 | 4 |
| Increase rural allowance and provide food | 1 | 2 | 1 | 4 | - | - | - | 0 | 4 |
| Recruit more health workers and ensure inexperienced staff are posted with experienced staff | 0 | 2 | 0 | 2 | - | 1 | - | 1 | 3 |
| Implement conflict resolution training and durbars to address community grievances | 1 | 0 | 0 | 1 | - | - | 2 | 2 | 3 |
| Teach women to ride motorbikes, equitable motorbike distribution and road improvements | 0 | 0 | 1 | 1 | - | 1 | - | 1 | 2 |
| Introduce security at facilities | - | - | - | - | - | 1 | 1 | 2 | 2 |
| Increase maternity leave and access to high quality schools for women HWs children | 0 | 1 | 0 | 1 | - | 1 | - | 1 | 2 |

## 4. Discussion

This study sought to elucidate the gendered HWF retention challenges in some of the most deprived Ghanaian districts and propose evidence-based recommendations for gender transformative actions. Overall, women dominate in facilities located in the major towns and communities around or within the deprived districts, with the few men posted on islands and remotest communities. In non-island districts, there is a shortage of women, primarily linked to romantic relationships/family, accommodation, transport and safety and security challenges. Regarding <u>access to resources</u>, women perceived accommodation, food /water and network and electricity as more important than men, and whilst both genders highlighted infrastructure/supplies and transport issues, women described unique transport challenges, including safety/security issues. In terms of <u>division of labour, roles and everyday practices</u>, most respondents highlighted that women bear the brunt of relationship and family responsibilities and although men perceived education for children as a key female attrition driver, women rarely mentioned it. Whilst women and men described mixed opinions of career progression, with negative perceptions in KE, there were gendered differences in <u>social norms and values</u>. Women commonly highlighted community conflict, financial challenges and unique safety and security challenges, including sexual harassment, with formal policies lacking. However, many women reported positive promotion and study leave experiences. Whilst men highlighted conflict between women HWs, it was rarely described by women. Throughout, respondents expressed stereotypical gender views, with women commonly perceived as less resilient, better suited nursing roles due to their empathy and caring nature and men deemed ‘fearless’. No respondents were aware of formal gender tailored policies; women’s gender transformative recommendations were primarily linked to accommodation, education, reducing motorbike use and improving financial incentives, whilst men focused on conflict and security.

### Attrition of women health workers is shaped by intersecting inequalities

Women HWs are experiencing intersecting inequalities, shaping relative power and experiences of living and working in deprived districts – we explore these using Simpson’s intersectionality wheel (25). The districts have low **socio-economic status**, directly contributing to challenges around basic amenities, including accommodation – a key attrition of women. Strong **postcolonial** cultural ideologies related to hegemonic masculinities – including men’s fearlessness and dominance – prevail in Ghana, driving women’s subordination, inequitable power and responsibility for domestic and caring roles (30–33). Research shows how these **patriarchal structures** are commonly mirrored and reinforced at the **health system** level, including the workforce, impeding UHC (34, 35). For instance, a review from Hay, McDougal (35) found that women are resigned to ‘caring’ roles as nurses and community health workers, whilst men dominate ‘curing’ roles as physicians or leadership positions (17). This perception was commonly highlighted in our findings and illustrated by the difficulties researchers faced identifying women in leadership positions to interview. Other studies in Ghana, including a recent evaluation by PwC (36), mirror our findings, with some women reportedly feeling that they must work harder than men to achieve managerial roles (37). Institutional policies and practices are also influenced by social norms and gender stereotypes, resulting in unresponsive policies, as illustrated by our study and discussed below (17). Ghana’s health system is chronically underfunded, driving insufficient infrastructure, supplies and staff shortages, highlighted by women HWs (38). This is driving attrition and further weakening the health system, echoing findings from another study in rural Ghana, where inadequate infrastructure drove attrition (39). These social dimensions interact with wider forms of discrimination and identity factors, shaping experiences of women HWs.

Women are based far away from their families and partners, with geographic location and gender norms intersecting with numerous factors, driving attrition. Intersecting challenges around **location**, **education**, **income** and **occupation** and feeling forced to choose between their identity as mothers and their careers, were commonly described. This was linked to the inability to conduct locum work, attend evening or weekend education/training courses due to the rural location, as well as insufficient financial incentives, creches and feeding rooms. Similar urban-rural professional development inequities were described in Nigeria (40) and among deprived Ghanaian nurses in another study, who reportedly experienced shame and a sense of “lagging behind” urban colleagues (41). Some reviews have elucidated societal beliefs that women should be driven by altruism rather than socioeconomic gain, which are mirrored in health systems, driving gender inequity, mirroring findings from our study (35, 42).

Whilst many women benefitted from the study leave and promotion policies, most men and women in KE did not. Moreover, almost all the men and women reported living far away from their partners, suggesting that the postings on marital grounds policy are not being fully considered. Women also highlighted a lack of creches and feeding rooms, and none of the respondents described receiving reduced working hours and one-hour feeding breaks upon returning to work, highlighting potential gaps in practice. This was echoed by another study that described a lack of creches in Ghanaian HFs (36). This suggests a gap between policy and practice, supporting findings from other studies on HWF policies in Ghana, which emphasise a lack of explicit procedures and awareness, resulting in unequal opportunities (43, 44). Overall, we found that the lack of familial support, expensive housing and cost of living, combined with the inadequate salary and lack of income generation opportunities, is hindering women’s socioeconomic independence.

Alongside this, women face societal pressures to find husbands, whilst partnered women bear responsibility for relationship maintenance and their children’s educational attainment, in the eyes of men. Consequently, many women are leaving to join their families, whilst men depart for “greener pastures”, echoing other studies in Ghana which found women typically leave due to marital and childbearing challenges (43–45). Due to socioeconomic dependence and relationship pressures, some women are refusing postings or relocating to join husbands, hindering career progression – as evidenced in other LMICs (46). Our findings align with global evidence, which asserts that women’s domestic and familial responsibilities, combined with unsupportive institutional policies and practice, drive occupational segregation, hindering women’s careers and directly benefitting men (17, 46–49).

**Physical health** also intersects with numerous factors, including geographic location and age. Community-level violence and sexual harassment were prevalent, with unemployment, insufficient security, unsafe housing and long outreach journeys being key determinants and isolation from support networks, stigma and a lack of policy awareness impeding reporting. Whilst evidence remains limited, some studies in Ghana report rates of workplace violence up to 68.2% (37) and 73.9% (50), with on-call duties and inexperience significantly increasing risk (37). Another qualitative Ghanaian study documented a high prevalence of sexual harassment among women HWs (51) and research shows that women located in remote rural settings conducting outreach are particularly at risk (17, 37, 52, 53), with young, unmarried midwives in Malawi and Ghana lacking secure accommodation reportedly worried about their safety – supporting our findings (54). Research shows how lower cadre staff have a higher risk of violence, with women HWs experiencing violence, including sexual harassment, if they are perceived as failing their role as caregiver, with gender norms of female passivity perpetuating violence (35, 37, 55, 56). This was reflected in our findings, wherein community members became violent when women refused to answer their doors late at night or took time off for annual leave or professional development. Crucially, we found a lack of awareness **of** formal policies on sexual harassment, with some respondents highlighting issues with stigma. This reflects findings from other studies in Ghana which report a lack of awareness and sufficient formal policies (37, 51). Moreover, whilst PwC (36) report that there is officially a zero tolerance policy, with sexual harassment sanctioned in the Code of Conduct and Disciplinary Procedures, there is a lack of awareness and training on sexual harassment and gender, including among the committees to which incidents are reported. Moreover, there is a degree of ambiguity as to what constitutes sexual harassment (36). Long motorbike journeys were commonly attributed to violence, worse injuries in pregnant women and miscarriages, with the latter also owed to psychological stress associated with distance from partners and difficult working conditions. Whilst the evidence base and quality is limited, a scoping review from Marsters, Stafl (57) suggests that HWs may have a heightened risk of miscarriages and preterm births, partially due to long working hours. Another review found that women previously exposed to psychological stress were significantly more likely to experience miscarriages, though literature was primarily from high income countries (OR 1.42, 95% CI 1.19–1.70) and a retrospective cohort study in Taiwan indicated pregnant women were more likely to experience severe motorcycle injuries (58, 59). Experiencing sexual harassment and motorcycle accidents is associated with physical and psychological burdens, which may drive attrition – especially in light of the lack of risk insurance and inadequate occupational health policies, described by respondents (37, 51, 60).

Overall, women HWs in deprived areas are being devalued, which is driving attrition. Aside from impeding the quality of care, systematic occupational segregation is influencing the career aspirations of future generations, as well as bestowing more power with “invisible gatekeepers”, primarily male managers and romantic partners, perpetuating inequities (35, 46). The ongoing retention crisis in rural Ghana threatens to exacerbate rural inequities and impede progress towards the SDGs, including UHC (61–63). Since the majority of Ghana’s HWF in deprived areas is women, and the equality and empowerment of the female HWF is central in achieving UHC, gender transformative interventions and policies are needed.

### Evidence-based recommendations for gender transformative action

Based on our findings, we suggest the following: the health system and community stakeholders must collaborate to provide appropriate, affordable and secure accommodation and improve facility security e.g., lighting and security personnel. To address sexual harassment, managers must actively promote awareness and reporting systems, as well as remaining vigilant. The health system should also conduct community engagement to improve knowledge, attitudes and practices surrounding sexual harassment and review sexual harassment policy to ensure that it is unambiguous and effective. Moreover, policy should be revised to prevent women from being posted on their own or in pairs with men only. To address the burden experienced by women with childcaring responsibilities, formal policies should be reviewed and or amended to provide mothers with increased financial allowances, access to creches and feeding rooms at work, as well as considerations when assigning annual leave, rota schedules and postings. Alongside this, the financial allowance for deprived HWs should be reviewed and or revised and the health system must garner support from communities in providing non-financial incentives, such as foodstuff and potentially support the implementation of informal saving schemes, such as a susu (41). Since appropriate implementation of study leave was a driver of retention and HWs emphasised that quotas were not being reached in some sub-districts, there should be an audit in deprived areas to ensure the policy is being implemented equitably. Moreover, the health system should review the occupational support and risk insurance for deprived HWs, and instate formal policy which prevents pregnant women from having to ride motorbikes and independent outreach by women. Finally, when co-designing health system strengthening interventions such as these, it is crucial that women are actively engaged throughout the research cycle and that interventions are context specific.

### Strengths and limitations

This study contributes to the limited evidence base and is informed by needs on the ground, expressed during participatory workshops. Findings have the potential to inform health system strengthening interventions and will be disseminated with Ghanaian MoH and key partners. Moreover, the study engaged informants with a range of experience and job roles, across the health system levels, enhancing trustworthiness and most interviews with women were conducted by female research team members. However, there are some limitations; firstly, most interviews with women were conducted several months later, when implementation of the community-based retention strengthening interventions had begun. Similarly, tools were amended (e.g., addition of questions related to sexual harassment and opportunities for gender tailoring) for the second round of interviews, potentially biasing results. To mitigate this, respondents were probed to consider the situation prior to the intervention, and transcripts from male respondents were screened for gender responsive recommendations. Additionally, we were unable to engage women HWs at senior health system levels, though this also reflects the reality of most women being concentrated at lower levels of the health system.

### Conclusion

The health workforce crisis in Ghana is threatening progress towards the SDGs, including UHC, especially in deprived districts where retention is lowest. Most HWs in deprived districts are women who face unique intersecting challenges which are shaping relative power and resilience, primarily related to accommodation, transport, safety and security and socioeconomic insecurity. Aside from impeding retention, these challenges are perpetuating gender inequities; therefore, gender transformative retention strategies are crucial in delivering equitable services and achieving UHC.

## Data Availability

Given that this study was conducted in a limited number of small sub-districts and the cadre and gender of respondents was disclosed for analysis purposes, making KII transcripts readily available may risk respondents being identified. This may have repercussions on their relationships with their employers, communities and partners, as they spoke openly about challenges faced. Moreover, some respondents disclosed very personal, sensitive experiences, including sexual violence. Therefore, we request that this study be exempt from making data publicly available.

## Acknowledgments

We would like to give thanks to all the respondents who took part in the study and to our funders, Global Health Partnerships.

